# Donor HLA class 1 evolutionary divergence is a major predictor of liver allograft rejection: a retrospective cohort study

**DOI:** 10.1101/2020.12.18.20245381

**Authors:** Cyrille Féray, Jean-Luc Taupin, Mylène Sebagh, Vincent Allain, Zeynep Demir, Marc-Antoine Allard, Christophe Desterke, Audrey Coilly, Faouzi Saliba, Eric Vibert, Daniel Azoulay, Catherine Guettier, Dominique Debray-Devictor, Sophie Caillat-Zucman, Didier Samuel

**Affiliations:** Centre Hépato-Biliaire, APHP, Hôpital Paul-Brousse, 94800 Villejuif, France. Université Paris-Saclay, UMR-S 1193 INSERM; FHU Hepatinov, 94805, Villejuif, France; Laboratoire d’Immunologie et Histocompatibilité, AP-HP, Hôpital Saint-Louis, 75010 Paris, France. Université de Paris, UMR 976 INSERM, 75010, Paris France; Laboratoire d’Anatomopathologie, APHP Hôpital Paul-Brousse, 94800, Villejuif, France. Université Paris-Saclay, UMR-S 1193 INSERM; Université Paris-Saclay, Physiopathogénèse et traitement des maladies du Foie; FHU Hepatinov, 94805, Villejuif, France; Unité d’hépatologie pédiatrique, APHP, Hôpital Necker Enfants Malades. Paris, Université de Paris. France

## Abstract

**Background:** Recognition of donor antigens by the recipient’s immune system leads to allograft rejection. HLA evolutionary divergence (HED) between an individual’s HLA alleles is a continuous metric that quantifies the differences between each amino acid of two homologous alleles and reflects the breadth of the immunopeptidome presented to T lymphocytes. We investigated whether or not HED of the donor or of the recipient has an impact on liver transplant rejection.

**Method:** We did a retrospective cohort study in 1154 adult and 113 children recipients of liver transplant. We considered the histological lesions in liver biopsies performed routinely 1,2 and 5 years after transplantation and in case of liver dysfunction. Donor-specific anti-HLA antibodies (DSA) were determined in children at the time of biopsy. HED was calculated using the physicochemical Grantham distance for class I (HLA-A, HLA-B) and class II (HLA-DRB1, HLA-DQB1) alleles. We assessed the incidence of rejection-related liver lesions using a multivariate Cox proportional hazards regression analysis.

**Findings:** In the adult cohort, recipients from donors with class I HED above the median had a higher risk of acute or chronic rejection, but not of other histological lesions. HED of the recipients was not related to any histological lesion. In multivariate analysis, a high donor class I HED was associated with acute rejection (hazard ratio [HR] 1.79; 95% confidence interval [CI]: 1.34-2.40; P<0.0001) or chronic rejection (HR 2.26, CI 1.45-3.51; P<0.0001) and was independent of age and HLA identities. In the pediatric cohort, class I HED of the donor was also associated with acute rejection (HR 1.81, 95% CI 1.12-3.14; P=0.013) independently of the presence of DSA.

**Interpretation:** Class I HED of the donor reflects graft immunogenicity and predicts rejection independently of donor-recipient HLA compatibility. This novel and easily accessible prognostic marker could improve donor selection and guide immunosuppression.

## Research in context

### Evidence before this study

Without immunosuppression, a liver transplant is rejected due to the recipient’s T-cell immune response to donor’s alloantigens. A low number of HLA identities between the recipient and the donor only weakly predicts liver allograft rejection, and HLA matching is usually not done in the liver transplantation setting. However, HLA mismatches may induce the production of donor-specific antibodies which could by themselves induce liver lesions.

T lymphocytes recognize non-self peptides presented within the peptide-binding groove of HLA class I and class II molecules. For each HLA locus, two codominant alleles are expressed and most patients are heterozygous. It has been demonstrated that the more divergent are homologous alleles, the broader is the immunopeptidome they present, and thus the stronger is the T-cell response. This HLA evolutionary divergence (HED) can be calculated as a distance metric evaluating the physicochemical differences between homologous amino acids composing the peptide-binding groove of two homologous alleles. HED was recently identified as the main driver of anti-tumor response after immune checkpoint inhibitor therapy in cancer patients. The role of HED has never been evaluated after organ transplantation, although it may have an impact on rejection.

### Added value of this study

To our knowledge, this study is the first to show that HED of the donor, an intrinsic parameter entirely independent of the characteristics of the recipient, is a major predictor of liver transplant rejection. A high class I HED of the donor was strongly related to the occurrence and severity of acute and chronic rejection, but not to other histological lesions. In contrast, HED of the recipient had no impact on rejection. This effect of donor HED was independent of other known predictors of rejection, in particular HLA mismatches between donor and recipient and the presence of donor-specific anti-HLA antibodies at the time of rejection.

### Implication of all the available evidence

The higher is the class I HED score of the donor (and therefore the broader the immunopeptidome presented to recipient’s T cells), the higher is the risk of rejection. HED of the donor is easy to calculate as soon as HLA typing has been done, and can be made available before transplantation. Recommendations might be proposed for accepting or not a graft from a donor according to his HED depending on the recipient rejection risk, or for modulating the required level of immunosuppression if there is no choice between several recipients.

## Introduction

Liver transplantation remains the only life-saving treatment for many patients with end-stage liver diseases. However, in the absence of immunosuppression, graft rejection is almost constant because of the recipient’s T and B cell responses to the donor’s HLA antigens^1,2^. An individual’s HLA genotype consists of a pair of alleles at each class I and class II locus, where polymorphism is concentrated in the exons that encode the peptide-binding groove of the HLA molecule. The divergent allele advantage hypothesis predicts that HLA genotypes with more divergent heterozygous alleles (i.e. a larger number of amino acid differences in peptide-binding domains) enable the presentation of a more diverse repertoire of peptides, referred to as the immunopeptidome, which in turn increases the probability of triggering a specific immune response^3^. For many years, the advantage of HLA allelic differences has been estimated by simply comparing homozygous and heterozygous subjects, and a strong advantage for heterozygosity has been observed in the control of viral diseases and the response to cancer immunotherapies ^4,5,6,7,8^.

More recently, HLA allele divergence was more precisely measured as a continuous metric using the Grantham distance^9^, which takes into account the differences in the composition, polarity and volume of each amino acid within the peptide-binding groove of two homologous HLA alleles, and has been found to better capture the functional properties of HLA molecules than other common distance metrics^3^. Experimental evidence confirmed that this so-called HLA evolutionary divergence (HED) between two homologous alleles was directly correlated with the total number of pathogen- or tumor-derived peptides bound by these two alleles^3,10^. Strikingly, HED emerged as a strong determinant of survival in cancer patients treated with immune checkpoint inhibitors, and in leukemia patients undergoing allogeneic hematopoietic stem cell transplantation^10,11^.

We hypothesized that HED might have an impact on the occurrence of rejection after liver transplantation, a situation where the donor and recipient are generally poorly HLA-matched given the marginal impact of HLA mismatching on the occurrence of liver rejection^12^. In such a context, a high HED score of the donor might increase the diversity of the graft-derived immunopeptidome targeted by the recipient’s T-cells, while a high HED score of the recipient might allow the recipient’s antigen-presenting cells to present a broader immunopeptidome to effector T cells. We therefore investigated whether HED of the donor or the recipient was correlated with the incidence of histological lesions of acute or chronic rejection.

## Methods

### Study design and participants

We conducted a retrospective study in two centers: Paul Brousse Hospital in Villejuif (adult cohort) and Necker-Enfants Malades Hospital in Paris (pediatric cohort). Characteristics of the patients and donors are shown in Table 1 and Supplementary Figure 1.

**Table 1.**
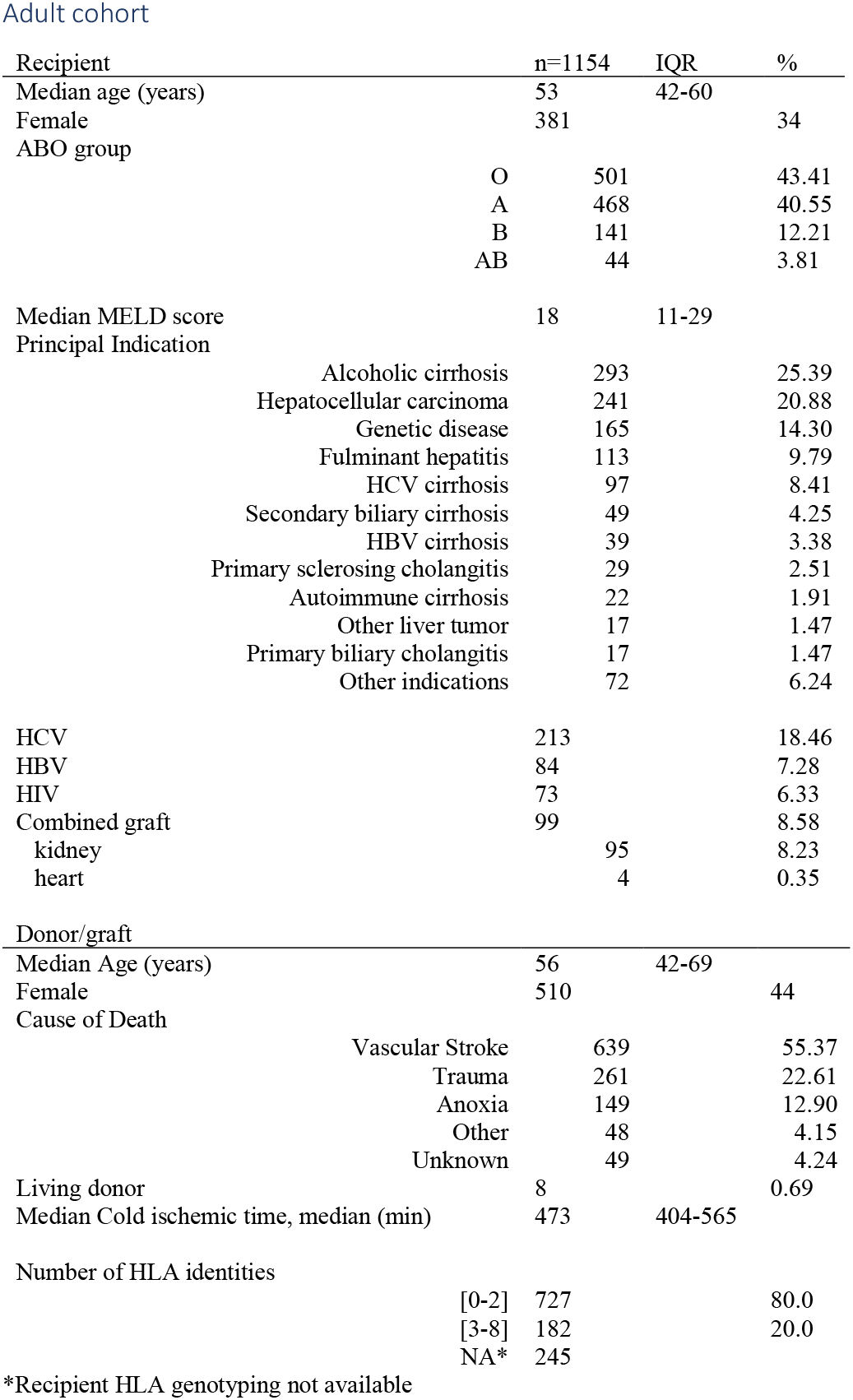
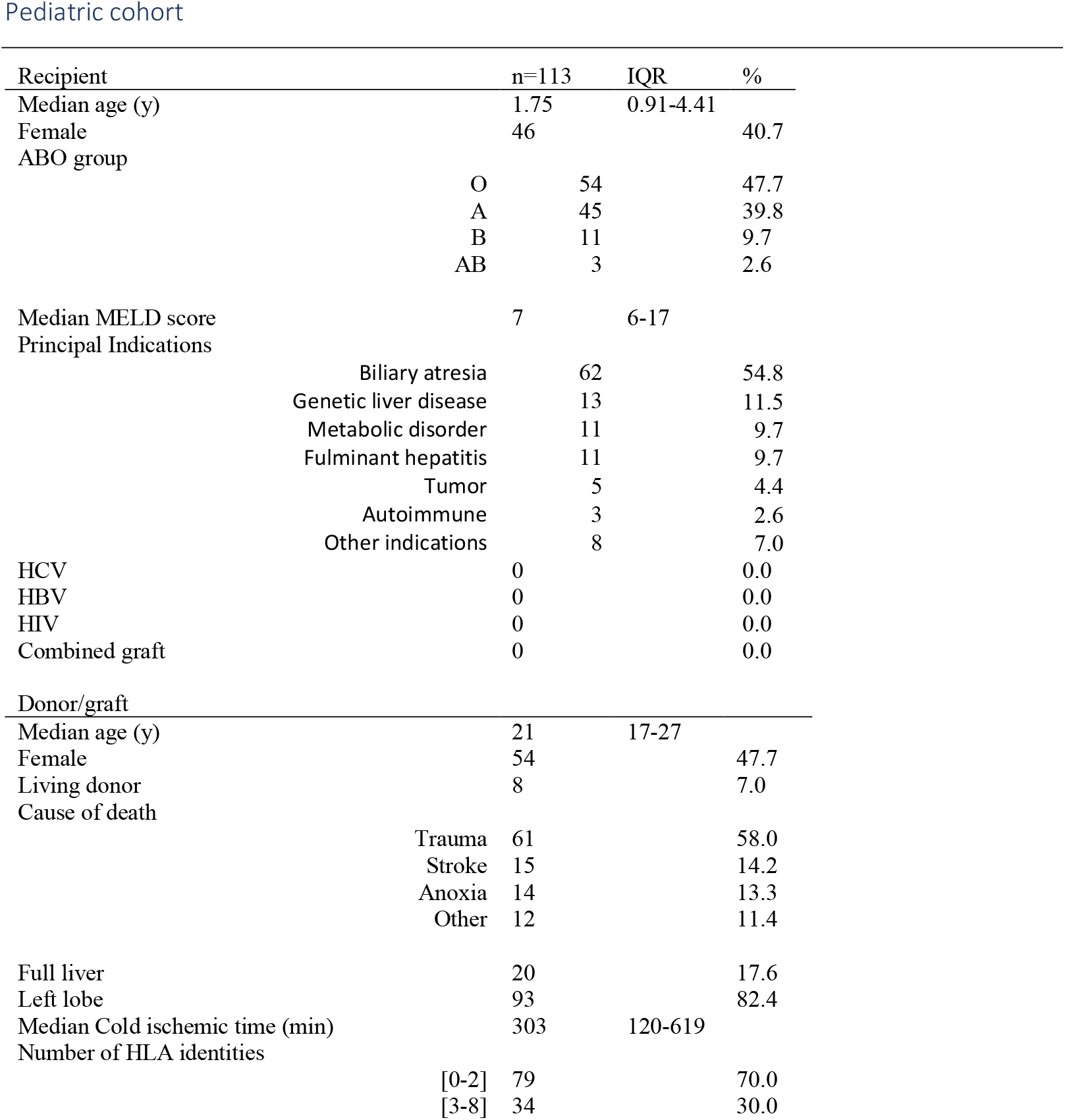
Recipient and donor characteristics in the adult and pediatric cohorts

Among the 1628 adult patients who received a first liver transplant between January 2004 and January 2018, donor HLA typing and post-transplant liver biopsy data were available for 1154 which constitute the adult cohort. Among them, recipient HLA typing data were available in 909 cases (79%). Donor-specific anti-HLA antibody (DSA) determination was not performed. Liver biopsies were performed after the first two weeks in the event of abnormal liver tests and no biliary obstruction. Routine biopsies were also systematically carried out at one, two and five years. Immunosuppression consisted of tacrolimus, tapered corticosteroids and mycophenolate. In the event of kidney- or heart-combined transplantation, the immunosuppressive regimen used anti-thymoglobulins, higher doses of immunosuppressive drugs and the maintenance of corticosteroids. HBV patients received hyperimmune anti-HBs polyclonal immunoglobulins^13^ with nucleos(t)ide anti-HBV analogue. All HIV patients were under anti-retroviral therapy. Before 2014, whenever possible, HCV patients were treated with pegylated interferon-alpha and ribavirin. From 2014, they received direct antiviral agents and were cured before or after transplantation.

Of the 172 children who received a first non-combined liver transplant between January 2010 and January 2018, 113 children alive at 01/01/2020 and for whom histological follow-up was available constituted the pediatric cohort. Routine biopsies were performed at one and five years. Donor and recipient HLA typing data were available for all patients. DSA were determined on the day of biopsy. Immune suppression was similar to that in adults, but the patients were weaned from corticosteroids during the first month post-transplant.

The study was carried out in accordance with French legal requirements and the Declaration of Helsinki.

### Procedures

Liver biopsy specimens were routinely paraffin-embedded and stained with hematein-eosin-safran and picrosirius. Throughout the study period, all liver biopsies performed in adult recipients were analyzed by two experienced pathologists (CG and MS) and reviewed with hepatologists during weekly staff meetings. For this study, we took into account the original diagnosis made at the time of biopsy: acute rejection, chronic rejection, ductopenia, steato-fibrosis, pattern of biliary obstruction, chronic hepatitis, cirrhosis, nodular regenerative hyperplasia, veno-occlusive disease and de novo immune hepatitis^14^. The Banff classifications were applied in case of acute rejection^15^. Early or late chronic rejection was based on the rates of ductopenia (20 to 50%, and ≥ 50%, respectively). In the pediatric cohort, the liver allograft fibrosis score was applied^16^.

HLA typing results were available at a first field (two-digit) resolution for the HLA-A, -B, -DRB1 and -DQB1 (but not HLA-C and -DPB1) loci. In recipients and living donors, HLA typing was performed using Luminex reverse polymerase chain reaction (PCR) sequence-specific oligonucleotides (PCR-SSO; One Lambda, Canoga Park, CA) with an increasing level of resolution over time. Deceased donors were typed using PCR sequence-specific primers (SSP; Olerup, Sweden until 2016, then Linkage Biosciences, CA). Complementary high resolution SSP typing of a donor was performed if DSA were detected after transplantation in any recipient having received an organ from this donor. Retyping was not performed during this study. Second field (four-digit) resolution typing was imputed from the most probable allele listed among ambiguities of the typing results. When more than one probable allele was proposed, we retained the most frequent allele based both on the description of the most frequent haplotypes encountered in the Paris region^17^ and with the HaploStats online tool (http://www.haplostats.org) run on the Caucasian and African populations which correspond to our recruitment. In our laboratory, such an imputation was found to be very consistent with high-resolution HLA genotyping using next-generation sequencing in over 1500 individuals (unpublished data). HLA matching was calculated for each recipient as the number of HLA-A, -B, -DRB1 and –DQB1 identities with the donor. Sequence divergence (at the amino acid level) between HLA alleles was computed for all possible combinations of allele pairs among alleles encountered in both cohorts for HLA-A, -B, -DRB1 and -DQB1 loci. The respective protein sequences of the peptide-binding groove (exons 2 and 3 for HLA class I, and exon 2 for HLA class II) were extracted from the IMGT/HLA database^18^. The calculation of HED between aligned allele pairs of a given locus was based on the Grantham distance metric^3^, a quantitative pairwise distance accounting for the physicochemical differences between two amino acids. For each subject and each donor, the mean class I HED and mean class II HED were calculated as the mean of the two pairwise divergences at the HLA-A and -B, and HLA-DRB1 and HLA-DQB1 loci, respectively, assuming that each locus contributes equally to the presentation of peptides^10^. By definition, there was a null divergence in case of homozygosity.

### Statistical analysis

As HED values are not normally distributed, nonparametric tests were used for comparisons. The cumulative incidences of each of the aforementioned liver graft lesions were calculated from the time of transplantation to the time of their first histological detection and compared between groups using the log-rank test for univariate analysis. The data were censored at the last available biopsy or at subsequent liver retransplantation. The multivariate Cox proportional hazards models were determined using predictors with a p-value <0.05 in univariate analysis. Risk was expressed as a hazard ratio with a 95% confidence interval using Cox’s regression. All statistical analyses were performed with R software 4.0 (R Foundation for Statistical Computing, Vienna, Austria; http://www.r-project.org, packages: ComplexHeatmap, survival, ggplot2).

## Results

We first performed a hierarchical clustering of HED for all pairwise allele combinations at the HLA-A, -B, -DRB1 and -DQB1 loci in donors and recipients in the adult and pediatric cohorts. Distinct clusters of high and low divergence between alleles were observed (Supplemental Figure 2). Pairwise divergences for HLA-B were higher than for HLA-A (p<0.0001), in line with previous reports that HLA-B is more diverse than HLA-A^10,19^. There was no correlation between class I or class II HEDs of donors and those of recipients, as expected because HED is an intrinsic parameter for each subject, regardless of donor or recipient status (Supplemental Table 1).

Among the 1154 adult recipients, 248 died without being re-transplanted, 65 were re-transplanted and 15 died after re-transplantation. The median follow-up period was 1464 days (IQR:707-2785). One- and five-year patient survival rates reached 93% and 80%, and one- and five-year graft survival rates were 92% and 76%. The median period elapsing between transplantation and the last available biopsy was 715 days (IQR 355-1826). During follow-up, the mean number of biopsies per patient was 2.9. In patients with normal liver function, the compliance to 1-, 2- and 5-year routine biopsies was 92 %, 56% and 85%, respectively. The last available biopsy was normal in 339 patients (29%). The most frequent histological lesions are shown in Table 2. The cumulative incidence of these lesions is shown in Supplemental Figure 3.

**Table 2.**
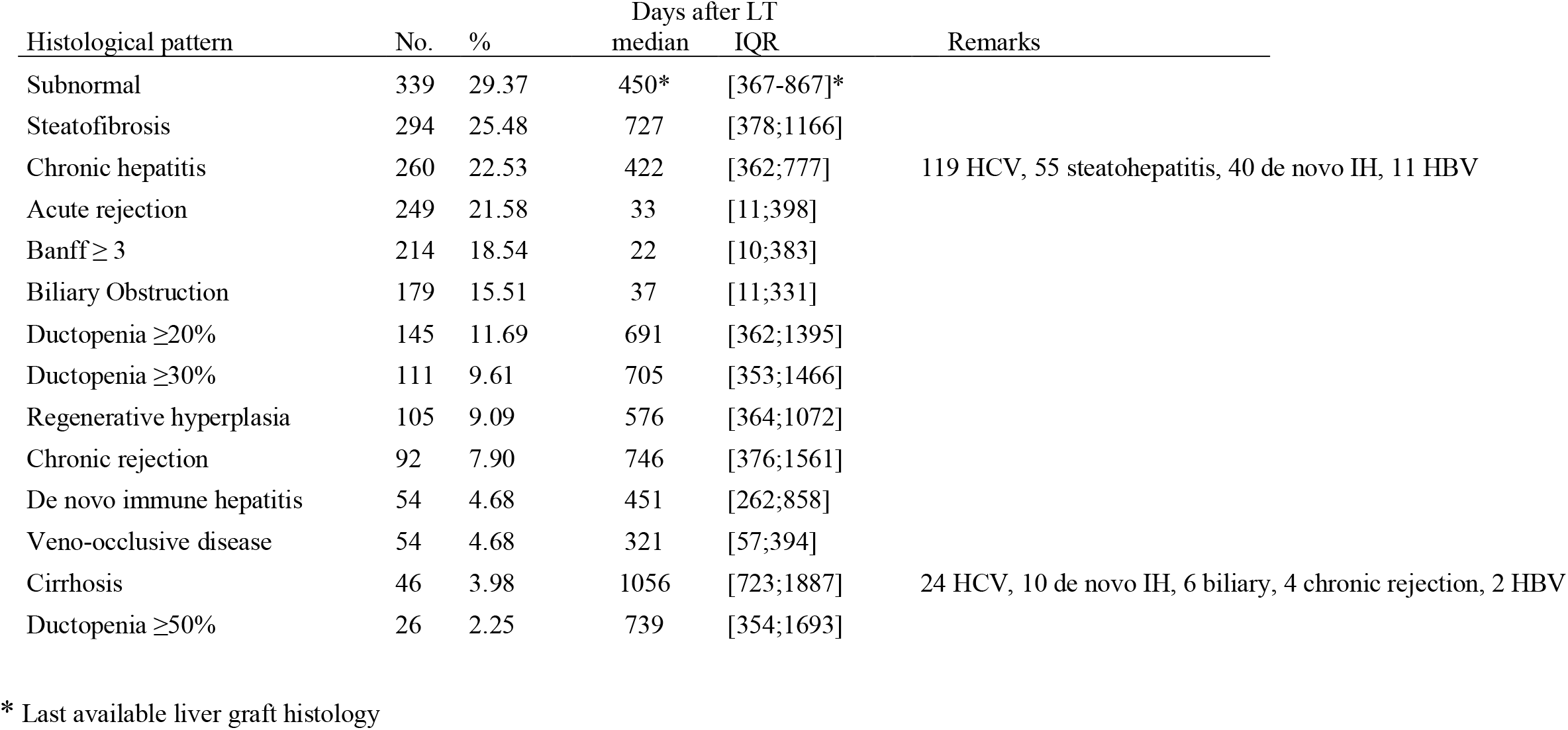
Post-transplant histological patterns in 1154 adult. For each lesion, the delay of detection was calculated from the time of liver transplantation (LT) to the first detection.

Donor or recipient class I and class II HEDs were not significantly related to donor or recipient variables such as age, gender, cause of liver transplantation, cause of donor death, liver steatosis in the donor, cold ischemia time and HCV, HIV or HBV infection, nor to the number of HLA incompatibilities or the CMV mismatch between donor and recipient (Supplemental Table 2). We correlated class I and class II HEDs of the donors or recipients with the cumulative incidence of each of the histological lesions mentioned above. Class I HED of the donors, expressed as continuous values or quartiles, were strongly related to rejection-related liver lesions such as acute or chronic rejection, acute rejection with Banff score ≥3, ductopenia, but not to any other lesions (Table 3). For acute rejection and acute rejection with Banff score ≥3, the top quartile cutoff threshold of donor class I HED demonstrated the most significant relationship. The five-year cumulative incidences of acute rejection were 40% and 22% in patients who received a liver transplant from a donor with class I HED above or below the top quartile, respectively (Figure 1A, P<0.0001). For chronic rejection and ductopenia, the median of class I HED values was the optimal cutoff threshold. For patients who received a transplant from a donor with a class I HED above the median, the risk of chronic rejection was 18% compared to 6% in those whose donor had a class I HED below the median (Figure 1B, P<0.0001). Among the 145 patients with ductopenia ≥ 20%, a strong positive relationship between the severity of ductopenia and class I HED of the donor was observed (Figure 1C). Analyses were repeated after exclusion of donors homozygous at HLA-A, HLA-B or both, leading to identical results. Importantly, class II HED of the donors, and class I and class II HEDs of the recipients were not related to any particular histological pattern (Supplemental Table 3).

**Figure 1:**
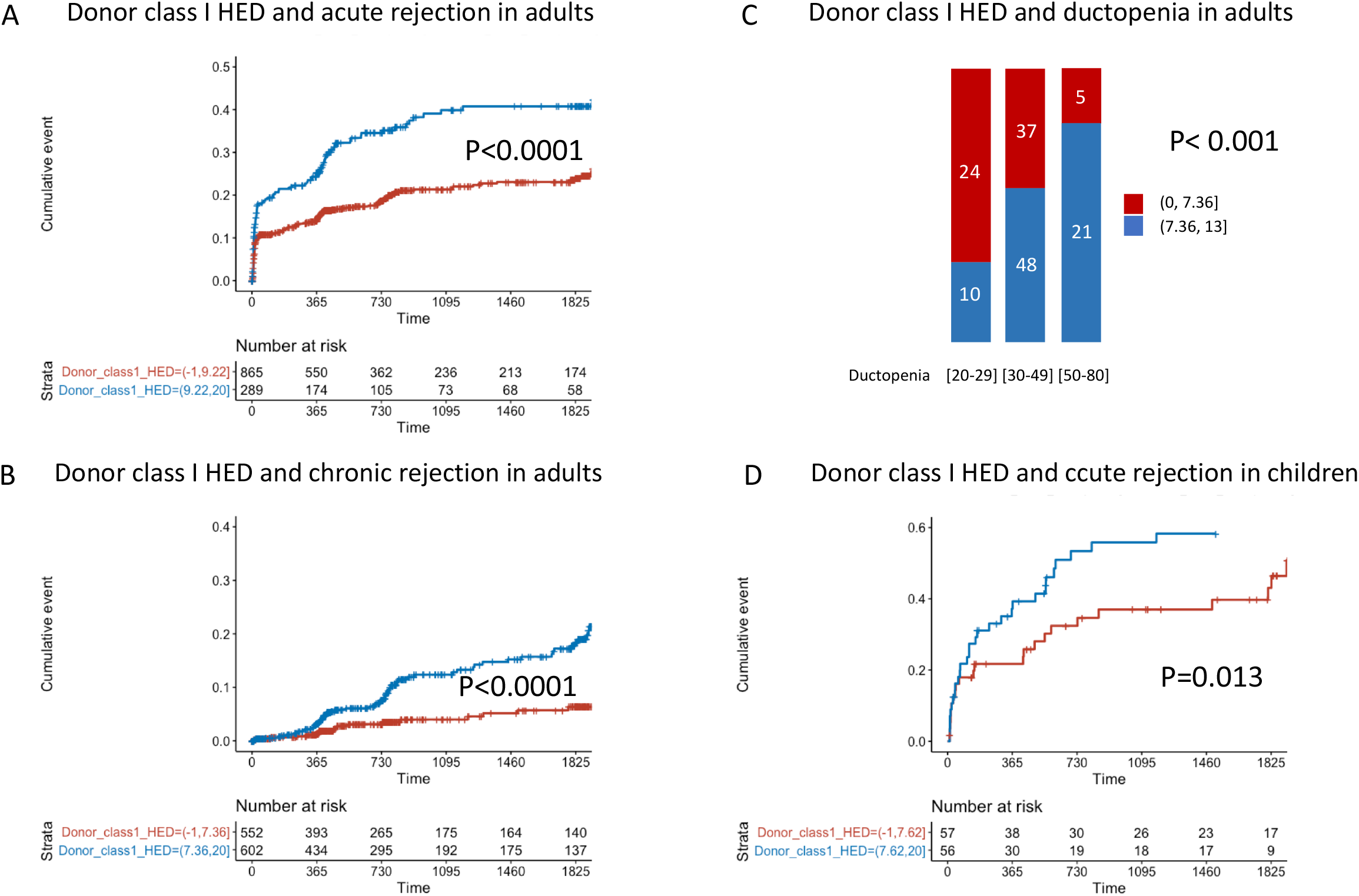
Effect of class I HED of the donor on histological lesions in liver transplant recipients. Panels A and B show the cumulative incidence of acute rejection (A) and chronic (B) rejection with regard to class I HED of the donor in the adult cohort. Panel C shows the relationship between the severity of ductopenia and donor class I HED (χ^2^== 23.96, p-value = 0.00053). Panel D shows the cumulative incidence of acute rejection in the pediatric cohort.

**Table 3.** Influence of donor class I HED on the cumulative incidence of histological lesions in the adult cohort (univariate analysis) Lower quartile of class I HED of the donor [0, 5.37] is used as reference and compared to other quartiles.

|  | Class I HED of the donor |  |  |  |  |  |  |  |  |  |
| --- | --- | --- | --- | --- | --- | --- | --- | --- | --- | --- |
|  | Second quartile<br>(5.37,7.41] |  |  | Third quartile<br>(7.41,9.22] |  |  | Top quartile<br>(9.22,20] |  |  | Log rank<br>p-value |
| Histology | HR | lower.95 | upper.95 | HR | lower.95 | upper.95 | HR | lower.95 | upper.95 |  |
| Steatosis | 0.6965 | 0.4987 | 0.9727 | 0.8318 | 0.5998 | 1.1536 | 0.8393 | 0.6114 | 1.1520 | 0.2 |
| Chronic hepatitis | 0.9724 | 0.6870 | 1.376 | 0.9242 | 0.6469 | 1.320 | 0.9750 | 0.6889 | 1.380 | 1.0 |
| **Acute rejection** | **0.8636** | **0.5751** | **1.297** | **1.2180** | **0.8325** | **1.782** | **1.7898** | **1.2569** | **2.549** | **0.0002** |
| **Banff ≥ 3** | **0.8713** | **0.5694** | **1.333** | **0.8713** | **0.5694** | **1.333** | **1.7727** | **1.2246** | **2.566** | **0.0001** |
| Biliary Obstruction | 0.8168 | 0.5250 | 1.271 | 0.9087 | 0.5893 | 1.401 | 1.3332 | 0.8975 | 1.981 | 0.08 |
| **Ductopenia ≥20%** | **0.5158** | **0.3055** | **0.8706** | **0.7108** | **0.4474** | **1.2391** | **0.9459** | **0.6220** | **1.6285** | **0.02** |
| Regenerative hyperplasia | 0.5961 | 0.3487 | 1.019 | 0.6158 | 0.3554 | 1.067 | 0.6440 | 0.3799 | 1.092 | 0.2 |
| **Chronic rejection** | **0.6264** | **0.2843** | **1.380** | **1.7949** | **0.9405** | **3.425** | **2.2805** | **1.2377** | **4.202** | **0.0002** |
| **Ductopenia ≥30%** | **0.3609** | **0.1875** | **0.6947** | **0.8450** | **0.5000** | **1.4280** | **1.1929** | **0.7429** | **1.9157** | **0.001** |
| De novo autoimmune disease | 0.7132 | 0.3392 | 1.500 | 0.6013 | 0.2700 | 1.339 | 0.9420 | 0.4703 | 1.887 | 0.5 |
| Veno-occlusive disease | 0.3463 | 0.1343 | 0.8929 | 1.0351 | 0.5168 | 2.0732 | 0.7868 | 0.3795 | 1.6310 | 0.09 |
| Cirrhosis | 1.065 | 0.4283 | 2.649 | 1.428 | 0.5825 | 3.502 | 1.554 | 0.6581 | 3.670 | 0.5 |
| **Ductopenia ≥50%** | **0.4049** | **0.0709** | **2.213** | **1.1544** | **0.3097** | **4.303** | **3.1217** | **1.0351** | **9.414** | **0.003** |

In univariate analysis, acute rejection was more common in case of a high class I HED of the donor (≥ 9.22 or top quartile), a small number of HLA antigenic identities between the donor and the recipient, HCV infection, HBsAg negativity, young recipient age (≤ 42 y), non-combined graft and autoimmune disease as indication of transplantation. In multivariate analysis, class I HED of the donor was the most significant variable associated with acute rejection (HR: 1.79; 95% CI: 1.34 to 2.40; P<0.001), followed by young recipient age, a non-combined graft and < 3 HLA antigenic identities (Figure 2, panel A).

**Figure 2:**
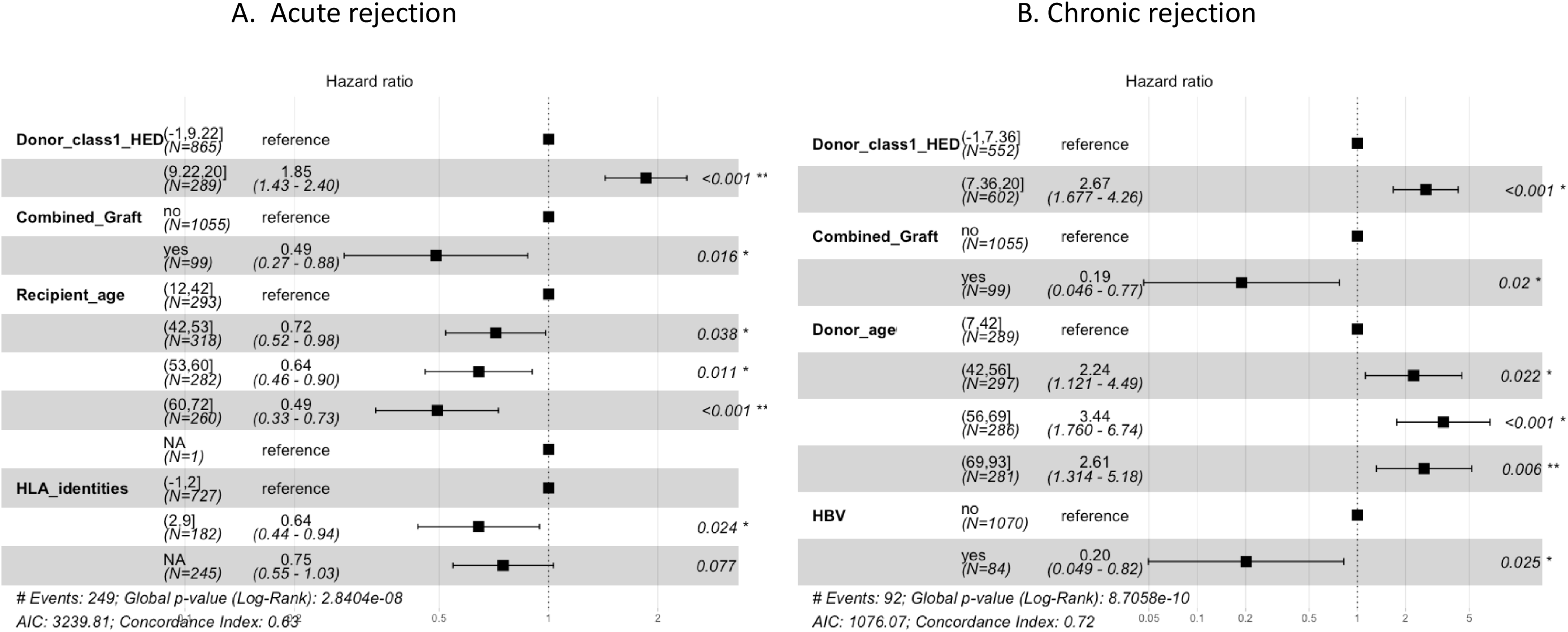
Multivariate Cox regression analysis of risk factors for acute and chronic rejection in the adult cohort. Estimated hazard ratios (with 95% confidence intervals) of acute rejection (A), and chronic rejection (B) in adult recipients.

Chronic rejection occurred more frequently in the event of a high donor class I HED (≥ 7.41 or median), HBsAg negativity, a donor older than 40 y, non-combined graft and an indication of fulminant hepatitis. In multivariate analysis, class I HED of the donor was the most significant variable (HR: 2.26; CI: 1.45 to 3.51; P<0.001), followed by donor age and HBsAg negativity (Figure 2, panel B).

We next used multivariate analysis to study other definitions of histological rejection. With respect to Banff-scored acute rejection (n=214), the model was very similar to that of acute rejection (n=249). When ductopenia ≥ 30% was considered as the endpoint of chronic rejection, only donor class I HED and donor age were significant. Finally, when ductopenia ≥ 50% was considered (n=30), it was only related to donor class I HED (Supplemental Figure 4).

Finally, we studied the role of HED in a distinct cohort of 113 liver transplant children. The median follow-up was 1668 days (IQR: 858-2521). Acute rejection and liver fibrosis were diagnosed in 63 (56%) and 61 (54%) children, respectively. The other histological lesions observed in adults were scarce: cirrhosis in two, steatosis in three, chronic rejection in four, de novo immune hepatitis in three. The two factors linked to acute rejection were the presence of class I DSA at time of biopsy (two-year cumulative incidence of rejection of 60 % in the 29 patients with positive class I DSA compared to 40% in the 84 others) and class I HED of the donor. The two-year cumulative incidence of acute rejection was 53 % when class I HED of the donor was above the median compared to 36 % when it was below this value (Figure 1, panel D; HR: 1.81; CI: 1.12 to 3.14, P=0.013). In multivariate analysis, both DSA and class I HED of the donor were significant and independent (Supplemental Figure 5). The presence of DSA was not correlated to class I or class II HED of the recipient or the donor. Liver fibrosis was not associated with any donor or recipient variables, including DSA and donor or recipient HED.

## Discussion

To our knowledge, this study is the first to show that HED of the donor, a divergence parameter calculated as an intrinsic metric for each donor individual, is an important predictor of allograft rejection after solid organ transplantation. The divergent allele advantage assumes that HLA genotypes with more divergent heterozygous alleles may allow the presentation of a larger set of peptides for T-cell recognition than less divergent alleles. Recent striking results in patients treated with anticancer immunotherapy^7,10^ highlighted the important role of class I HED in shaping the immunopeptidome and thus enhancing the antitumor response. Through this latter finding, not only has the HLA divergence hypothesis been reinforced, but a direct clinical application is offered by this metric of immunogenicity.

We showed here that a high class I (HLA-A and -B) HED of the donor is a strong and independent driver of acute and chronic rejection in a large, single-center cohort of adult liver transplant recipients with long-term follow-up and systematic and diagnostic liver biopsies. HLA-C typing was not available in our study, as it is not required for organ transplantation. However as recently demonstrated, HLA-C contributes much less than HLA-A and -B to the mean class I HED because of less divergence between HLA-C alleles^10,11^. The effect of class I HED of the donor on acute rejection was replicated in a smaller, independent pediatric cohort. Notably, class I or class II HED of the recipient had no impact on rejection.

These data suggest that the more divergent HLA class I molecules of the donor are, the more diverse are the graft-derived peptides they present to the recipient’s cytotoxic T-cells, and the higher the risk of rejection. Conversely, an increase in the diversity of the peptides presented by more divergent HLA alleles on the recipient’s APCs is not predictive of liver allograft rejection. Thus, what seems important is the breadth of the repertoire of (allogeneic) peptides bound to donor HLA alleles, and not the ability of recipient HLA alleles to bind a more diverse (self or allogeneic) repertoire. This principle is consistent with the marginal impact of donor-recipient HLA matching on liver allograft survival which has been largely described by others^12^ and was confirmed in the present study. Indeed, a low number of HLA identities with the donor was a weak predictor of acute rejection and was independent of HED.

Donor-specific HLA antibodies are mostly produced through the indirect presentation of allogeneic epitopes by recipient HLA class II molecules, providing CD4 T-cell help for the generation of antibody-producing B cells. Because the incidence of antibody-mediated rejection is low and this issue still debated in the liver transplantation setting, DSA determination has only recently been performed on a routine basis^15^. Therefore, we could not determine if HED is associated with the development of a humoral response to the graft in the adult cohort. In children tested for DSA at the time of liver biopsy, the presence of class I DSA was associated with acute rejection independent of class I HED of the donor. However, the presence of DSA was not related to HED of the donor or recipient. It will be of great interest to further evaluate the relationship between HED and occurrence of DSA in prospective studies in liver transplantation, but also in other solid organ transplantation settings (kidney, lung, heart) where the humoral response is crucial.

As well as HED, other known variables were involved in the risk of rejection. Recipient age was determinant in the case of acute rejection, as recently described by one of the few centers implementing a policy of routine biopsies^20^. This effect could be explained by both compliance issues in young recipients and so-called immune senescence ^21^ in older recipients. Rejection was less frequent in patients undergoing combined transplantation in whom immunosuppression was stronger than with liver transplantation alone. We also found less histological evidence of acute rejection in HBV patients, as we had observed previously^22^, and we confirmed our earlier findings on ductopenia and donor age^23^.

From a practical standpoint, class I HED of the donor was found to be more important than any other predictors of rejection. HED can be determined rapidly, at no additional cost as soon as the HLA genotype of the donor is available, regardless of the potential recipients. HED scores could be provided for large panels of combinations of common well-documented HLA class I alleles and made available for transplant centers once for ever as this parameter will not vary between any pair of alleles. This would offer opportunities for better selection of the donor in recipients with a higher risk of rejection. In adults, liver rejection seems to be controlled by immunosuppressive drugs. However, liver transplants with normal histology are infrequent^24,25^ and long-term prognosis is a major issue, especially in young recipients^26^. Selecting donors with low HED values for young recipients should enable better long-term histological results. In addition, HED could also be considered in personalized strategies to optimize immunosuppression as a function of the risk of rejection^27^.

## Data Availability

All the data are available

---

Supplemental Figure 1

Flowchart of the adult and pediatric patient cohorts

Supplemental Figure 2: Distribution of HLA evolutionary divergence (HED) at HLA-A, HLA-B, HLA-DRB1 and HLA-DQB1 in the adult and pediatric cohorts.

Panel A shows hierarchical clustering of HED at the different loci. At a given HLA locus, the heatmap shows HED values for all combinations across all alleles in all subjects. The color gradient represents HED values between allele pairs from low values (blue) to high values (red). Panel B shows violin plots of donor and recipient HEDs at each HLA locus. Null values of HED correspond to homozygous allele combinations.

Supplemental Figure 3

Cumulative incidence of histological lesions in adult liver transplant recipients.

Supplemental Figure 4

Multivariate Cox regression analysis of risk factors for Banff-scored acute rejection, ductopenia ≥ 30 % and ductopenia ≥ 50% in the adult cohort.

Supplemental Figure 5

Multivariate Cox regression analysis of risk factors for acute rejection in the pediatric cohort of liver transplant recipients (113 children).

