## Supplemental for "Donor HLA class 1 evolutionary divergence is a major predictor of liver allograft rejection: a retrospective cohort study"

### Supplemental Figure 1

Flowchart of adult and pediatric patients

Supplemental figure 1

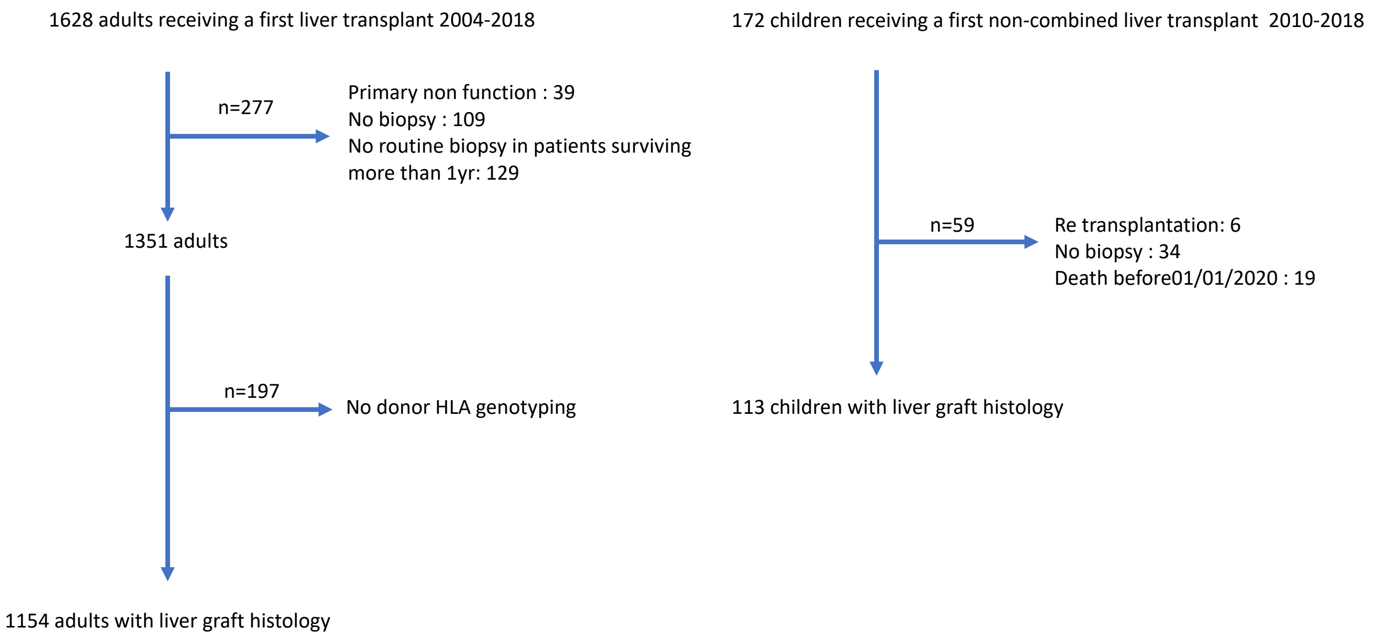

### Supplemental Figure 2: Distribution of HLA evolutionary divergence (HED) at HLA-A, HLA-B, HLA-DRB1 and HLA-DQB1 in the adult and pediatric cohorts.

Panel A shows hierarchical clustering of HED at the different loci. The heatmap shows HED values across all alleles in all subjects. The color gradient from blue to red indicates low HED values between allele pairs to high HED values between allele pairs, respectively. Panel B shows violin plots of donor and recipient HEDs at each HLA locus. Null values of HED corresponds to homozygous allele combination.

Supplemental figure 2A

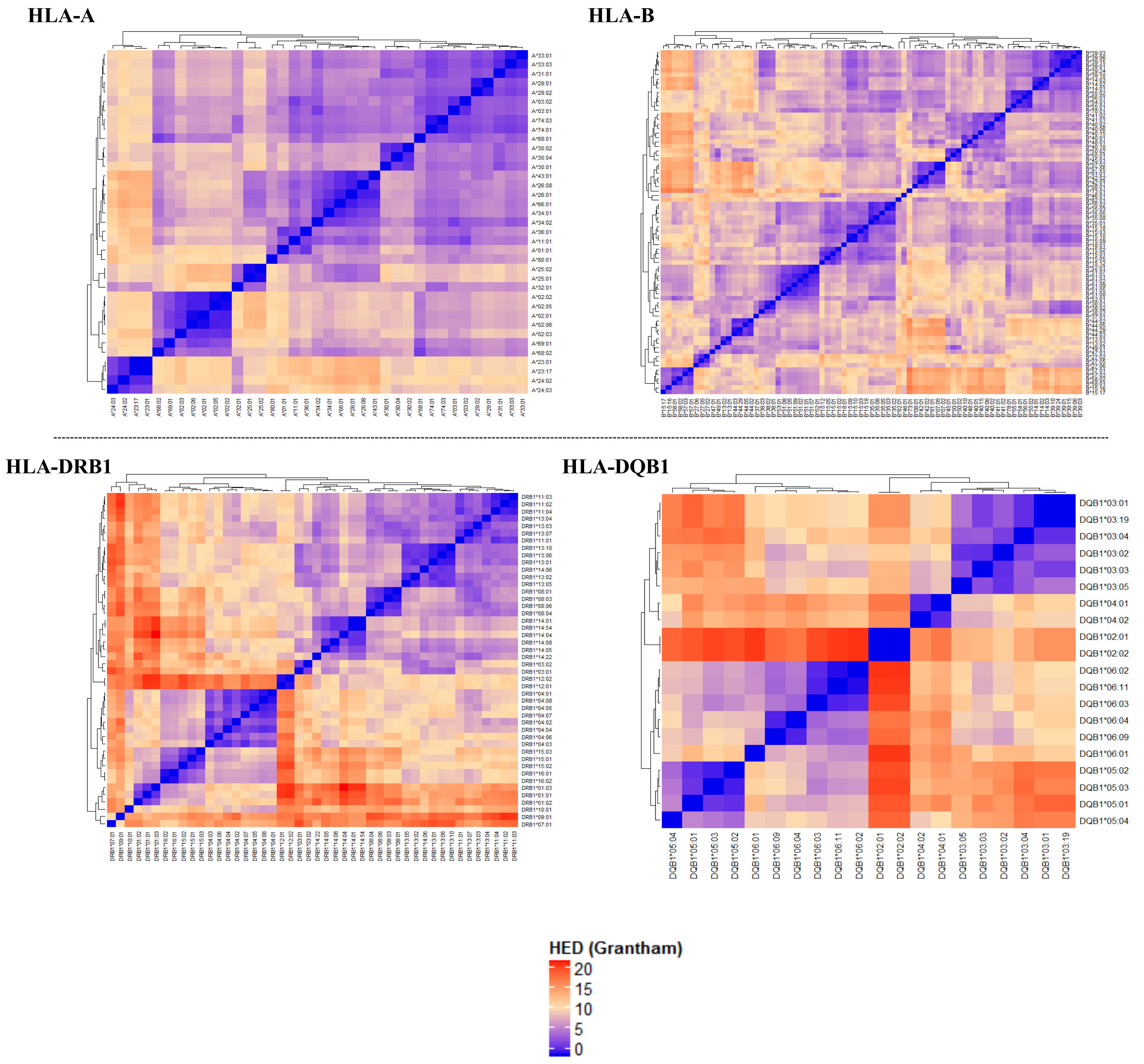

Supplemental figure 2B

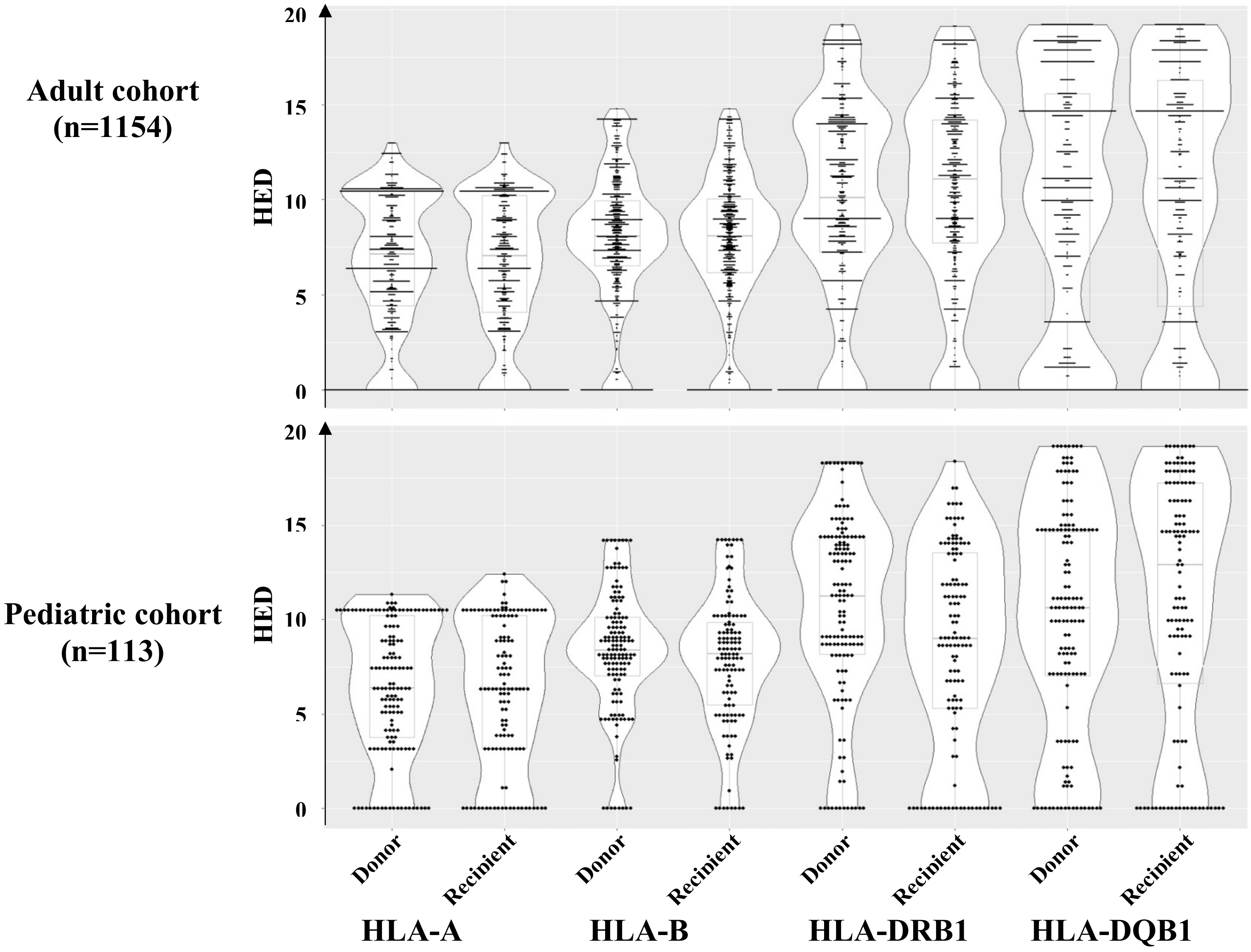

### Supplemental Figure 3

Cumulative incidence of histological lesions in adults.

**Supplemental figure 3**

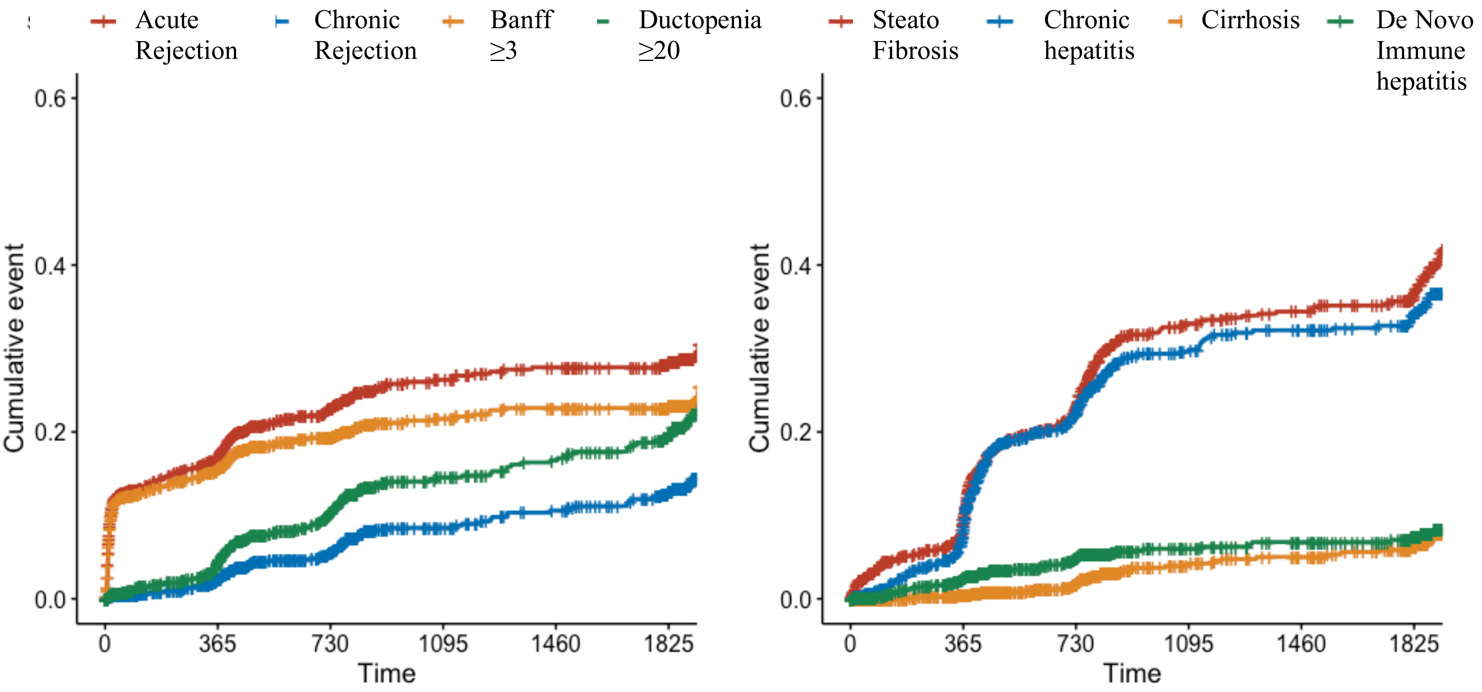

### Supplemental Figure 4

Multivariate Cox regression analysis of risk factors for Banff-scored acute rejection, ductopenia ≥ 30 % and ductopenia ≥ 50% in the adult cohort.

A. Banff ≥ 3

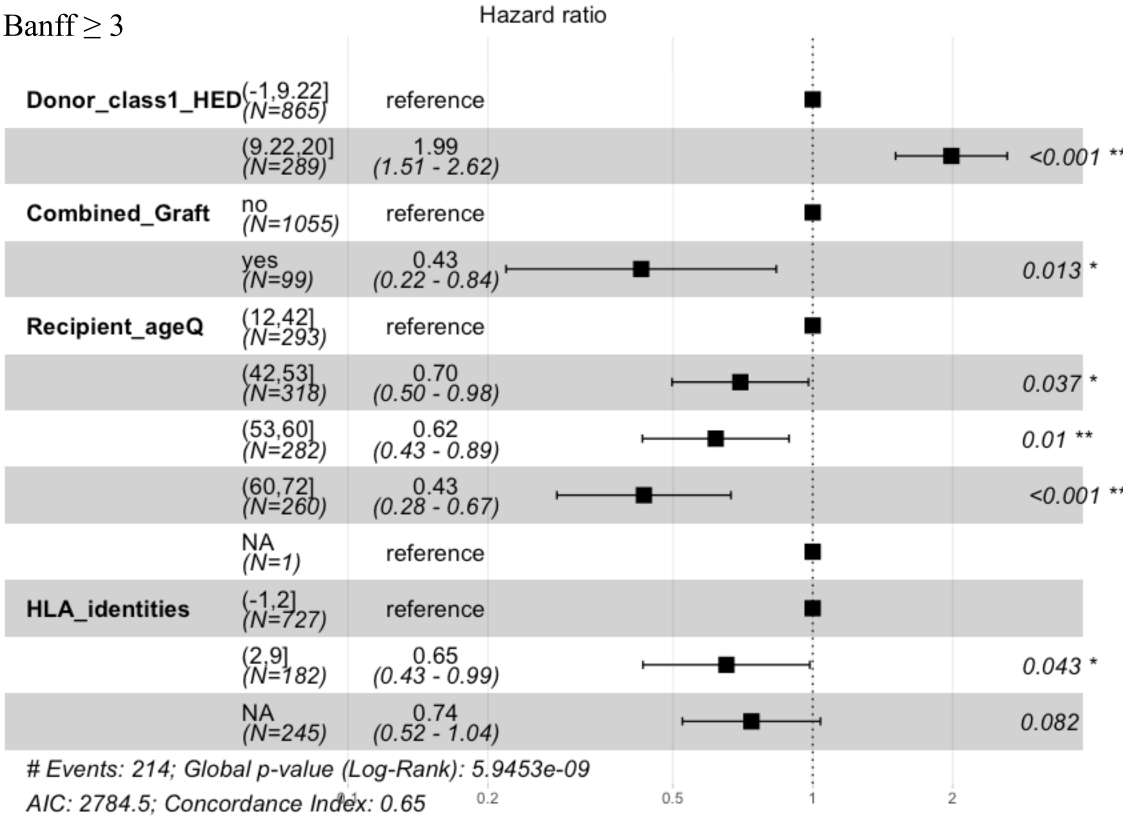

B. Ductopenia ≥ 30%

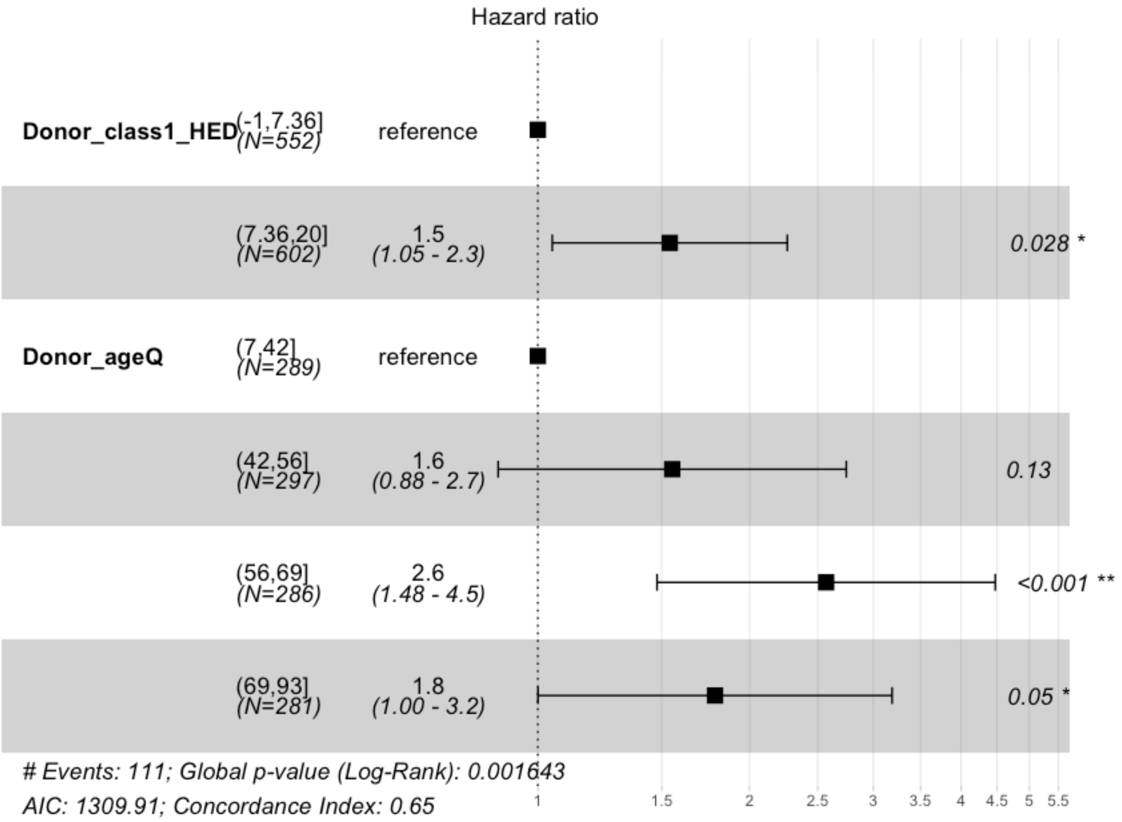


C. Ductopenia ≥ 50%

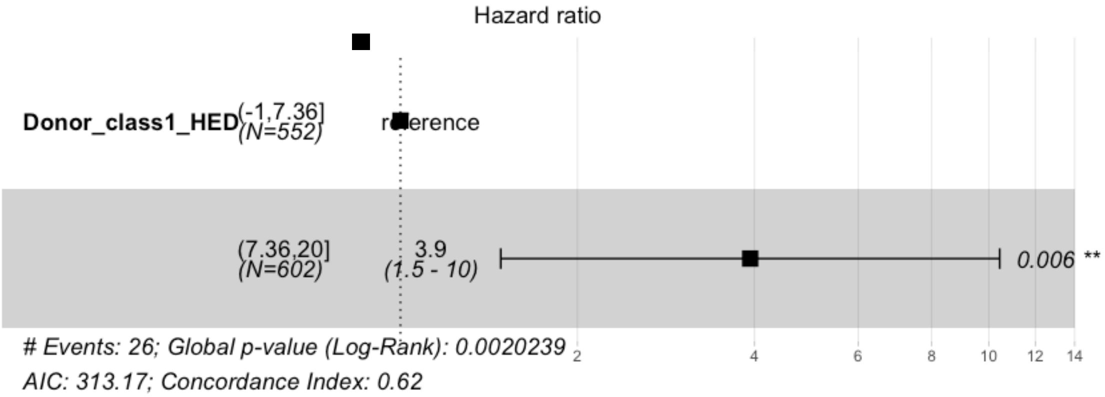

Supplemental Figure 5

Multivariate Cox regression analysis of risk factors for acute rejection in the pediatric cohort of liver transplant recipients (113 children).

Supplemental figure 5

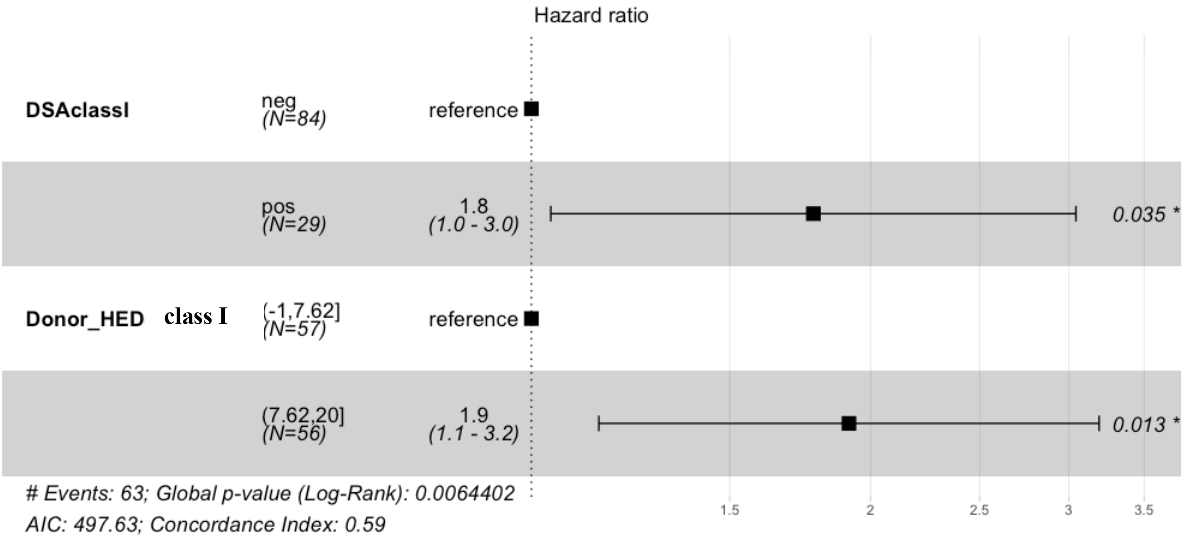

### Supplemental Table 1

Correlation between donor and recipient mean class I and class II HEDs (Kendall's rank correlation)

| | Donor class II HED | Recipient class I HED | Recipient class II HED |
|---|---|---|---|
| Donor class I HED | Tau=0.006, p-value = 0.771 | Tau=-0.007 ; p-value = 0.749 | Tau=0.008, p-value = 0.693 |
| Donor class II HED | - | Tau=0.013, p-value = 0.472 | Tau=-0.013 ; p-value = 0.6274 |
| Recipient class I HED | - | - | Tau=0.016 ; , p-value = 0.4557 |
| Recipient class II HED | - | - | - |

### Supplemental Table 2

Relationship between donor and recipient class I and II HEDs and main recipient and donor variables (Wilcoxon-Mann-Whitney test)

| | | Donor | | | | Recipient | | | |
|---|---|---|---|---|---|---|---|---|---|
| | | HED class I median[IQR] | p-value | HED class II median[IQR] | p-value | HED class I median[IQR] | p-value | HED class II median[IQR] | p-value |
| Recipient | | | | | | | | | |
| HIV positive | yes | 7.27 [5.69;9.25] | 0.46 | 10.44 [5.28;12.10] | 0.08 | 7.62 [5.25;9.18] | 0.94 | 11.71 [8.04;13.85] | 0.75 |
| | no | 7.52 [5.20;9.51] | | 10.87 [7.55;14.04] | | 7.51 [5.49;8.97] | | 11.06 [7.91;14.61] | |
| HBV positive | yes | 7.47 [5.97;9.98] | 0.80 | 10.10 [8.51;12.87] | 0.34 | 7.76 [5.70;9.00] | 0.72 | 10.90 [7.78;13.87] | 0.49 |
| | no | 7.52 [5.18;9.49] | | 10.90 [7.57;13.88] | | 7.50 [5.47;8.99] | | 11.10 [7.95;14.67] | |
| HCV positive | yes | 7.25 [5.67;9.23] | 0.63 | 10.55 [6.06;12.94] | 0.06 | 7.57 [5.43;9.16] | 0.70 | 11.31 [8.21;15.06] | 0.16 |
| | no | 7.55 [5.66;9.02] | | 10.89 [7.96;14.04] | | 7.51 [5.52;8.97] | | 10.98 [7.80;14.37] | |
| Gender | male | 7.32 [5.14;9.50] | 0.39 | 10.91 [7.12;13.63] | 0.61 | 7.51 [5.25;9.09] | 0.84 | 10.42 [8.12;14.38] | 0.86 |
| | female | 7.39 [5.81;9.65] | | 10.89 [7.45;14.07] | | 7.51 [5.69;8.97] | | 11.25 [7.80;14.71] | |
| Autoimmune | yes | 7.37 [5.32;9.17] | 0.58 | 11.28 [7.80;13.73] | 0.58 | 7.51 [5.45;8.99] | 0.65 | 10.90 [8.59;14.96] | 0.90 |
| | no | 7.53 [5.20;9.50] | | 10.89 [7.12;14.56] | | 7.52 [5.97; 9.22] | | 11.10 [7.87;14.56] | |
| Age | | | | | | | | | |
| | ≤ 53 y | 7.50 [5.56;9.22] | 0.54 | 10.90 [7.12;13.83] | 0.52 | 7.57 [5.82;9.11] | 0.11 | 11.33[7.94;14.38] | 0.90 |
| | >53 y | 7.52 [5.81;9.15] | | 10.08 [9.49;13.72] | | 7.33 [5.29;8.94] | | 10.33[7.91;14.70] | |
| Donor | | | | | | | | | |
| Gender | male | 7.43 [5.57;9.94] | 0.12 | 10.90 [7.12;14.10] | 0.54 | 7.43 [5.47;8.94] | 0.17 | 11.07[7.73;14.49] | 0.28 |
| | female | 7.57 [5.77;9.32] | | 10.89 [7.26;13.65] | | 7.56 [5.51;9.12] | | 11.16 [8.16;14.62] | |
| Age | | | | | | | | | |
| | ≤56 y | 7.52 [5.74;9.26] | 0.94 | 10.90 [7.12;14.10] | 0.34 | 7.45 [5.38;5.69] | 0.47 | 11.10[7.92;14.70] | 0.96 |

| | | | | | | | | | |
|---|---|---|---|---|---|---|---|---|---|
| | >56 y | 7.51 [5.83;9.10] | | 10.89 [7.55;13.30] | | 7.54 [5.69;8.99] | | 11.06[7.94;14.49] | |
| HLA identities | | | | | | | | | |
| | ≤2 | 7.64 [5.63;9.10] | 0.58 | 10.89 [7.75;13.30] | 0.09 | 7.57 [5.52;9.00] | 0.29 | 11.10 [7.89;14.68] | 0.58 |
| | >2 | 7.51 [5.93;9.34] | | 11.59 [8.12;14.70] | | 7.33 [5.18;8.93] | | 10.90 [8.12;14.03] | |
| CMV matching | | | | | | | | | |
| | D+/R- | 7.38 [5.35;9.08] | 0.58 | 10.86 [7.02;13.65] | 0.58 | 7.34 [5.29;9.07] | 0.89 | 10.68 [7.82;14.12] | 0.19 |
| | others | 7.25 [5.72;9.19] | | 11.28 [7.95;14.04] | | 7.54 [5.52;8.99] | | 11.28 [8.12;14.71] | |

### Supplemental Table 3:

Relationship between donor and recipient mean class I and II HEDs and liver histological lesions (univariate analysis) in adults

Lower quartile as reference

DONOR MEAN CLASS II HED

| | (5.41,7.87] | | | (7.87, 10.6] | | | (10.6,20) | | | Logrank p-value |
|---|---|---|---|---|---|---|---|---|---|---|
| Histology | HR | lower.95 | upper.95 | HR | lower.95 | upper.95 | HR | lower.95 | upper.95 | |
| Steatosis | 0.8761 | 0.6173 | 1.243 | 1.1939 | 0.8675 | 1.643 | 1.2530 | 0.9124 | 1.721 | 0.1 |
| Chronic Hepatitis | 1.0704 | 0.7525 | 1.523 | 1.0704 | 0.7525 | 1.523 | 1.1175 | 0.7971 | 1.567 | 0.9 |
| Acute Rejection | 0.8474 | 0.5905 | 1.216 | 1.0236 | 0.7339 | 1.428 | 0.8456 | 0.5957 | 1.200 | 0.6 |
| Banff ≥ 3 | 0.8179 | 0.5564 | 1.202 | 0.9709 | 0.6807 | 1.385 | 0.7596 | 0.5193 | 1.111 | 0.4 |
| Biliary Obstruction | 1.0021 | 0.6496 | 1.546 | 1.4362 | 0.9745 | 2.117 | 0.7732 | 0.4931 | 1.212 | 0.08 |
| Ductopenia ≥30% | 1.4012 | 0.8920 | 2.201 | 1.2790 | 0.8182 | 1.999 | 0.8102 | 0.4927 | 1.332 | 0.1 |
| Regenerative Hyperplasia | 0.6901 | 0.3750 | 1.270 | 1.1937 | 0.7148 | 1.994 | 0.9800 | 0.5757 | 1.668 | 0.3 |
| Chronic Rejection | 1.1376 | 0.6338 | 2.042 | 1.2736 | 0.7331 | 2.213 | 0.8389 | 0.4568 | 1.541 | 0.5 |
| De Novo Immune Disease | 0.7092 | 0.3217 | 1.563 | 1.0678 | 0.5392 | 2.115 | 0.8880 | 0.4333 | 1.820 | 0.8 |
| Veno-Occlusive Disease | 0.6881 | 0.3122 | 1.516 | 0.9104 | 0.4500 | 1.842 | 0.7356 | 0.3480 | 1.555 | 0.8 |
| Cirrhosis | 0.8754 | 0.3886 | 1.972 | 0.8767 | 0.3973 | 1.934 | 0.8342 | 0.3786 | 1.838 | 0.7 |
| Ductopenia ≥50% | 0.9866 | 0.4084 | 2.384 | 0.5985 | 0.2208 | 1.622 | 0.3820 | 0.1216 | 1.200 | 0.3 |

RECIPIENT MEAN CLASS I HED

| | (5.43, 7.51] | | | (7.51,9.03] | | | (9.03,20] | | | logrank p-value |
|---|---|---|---|---|---|---|---|---|---|---|
| Histology | HR | lower.95 | upper.95 | HR | lower.95 | upper.95 | HR | lower.95 | upper.95 | |
| Steatosis | 0.8573 | 0.6026 | 1.220 | 0.9657 | 0.6761 | 1.379 | 1.0573 | 0.7508 | 1.489 | 0.7 |

| | | | | | | | | | | |
|---|---|---|---|---|---|---|---|---|---|---|
| Chronic Hepatitis | 0.8898 | 0.6099 | 1.298 | 0.8095 | 0.5492 | 1.193 | 1.0930 | 0.7645 | 1.563 | 0.4 |
| Acute Rejection | 0.8700 | 0.5943 | 1.274 | 1.0237 | 0.7055 | 1.485 | 0.9761 | 0.6749 | 1.412 | 0.8 |
| Banff ≥ 3 | 0.7596 | 0.5193 | 1.111 | 0.9472 | 0.6347 | 1.414 | 0.9472 | 0.6347 | 1.414 | 0.6 |
| Biliary Obstruction | 0.7312 | 0.4743 | 1.127 | 0.8026 | 0.5222 | 1.233 | 0.6802 | 0.4366 | 1.060 | 0.3 |
| Ductopenia ≥30% | 0.8102 | 0.4927 | 1.332 | 0.8269 | 0.4910 | 1.393 | 1.1691 | 0.7293 | 1.874 | 0.6 |
| Regenerative Hyperplasia | 0.7792 | 0.4352 | 1.395 | 1.1753 | 0.6870 | 2.011 | 0.8033 | 0.4495 | 1.436 | 0.4 |
| Chronic Rejection | 0.7810 | 0.4289 | 1.422 | 0.6466 | 0.3374 | 1.239 | 1.0638 | 0.6097 | 1.856 | 0.4 |
| De Novo Auto Immune Disease | 1.4118 | 0.6850 | 2.910 | 0.7462 | 0.3143 | 1.771 | 0.8369 | 0.3691 | 1.897 | 0.3 |
| Veno-Occlusive Disease | 1.6459 | 0.6904 | 3.924 | 1.6415 | 0.6803 | 3.961 | 0.9369 | 0.3516 | 2.496 | 0.4 |
| Cirrhosis | 1.088 | 0.4505 | 2.626 | 1.026 | 0.4070 | 2.586 | 1.273 | 0.5363 | 3.023 | 0.9 |
| Ductopenia ≥50% | 0.7848 | 0.2637 | 2.336 | 0.7258 | 0.2303 | 2.288 | 1.3559 | 0.5161 | 3.562 | 0.6 |

RECIPIENT MEAN CLASS II HED

| | (5.869,8.046] | | | (8.046, 10.9] | | | (10.9,20] | | | logrank p-value |
|---|---|---|---|---|---|---|---|---|---|---|
| Histology | HR | lower.95 | upper.95 | HR | lower.95 | upper.95 | HR | lower.95 | upper.95 | |
| Steatosis | 0.6431 | 0.4452 | 0.9289 | 0.7937 | 0.5637 | 1.1176 | 0.7233 | 0.5088 | 1.0281 | 0.09 |
| Chronic Hepatitis | 0.882 | 0.5713 | 1.362 | 1.311 | 0.8833 | 1.946 | 1.311 | 0.8833 | 1.946 | 0.2 |
| Acute Rejection | 0.8588 | 0.5762 | 1.280 | 0.8339 | 0.5620 | 1.237 | 1.0893 | 0.7513 | 1.579 | 0.5 |
| Banff ≥ 3 | 0.9191 | 0.6035 | 1.400 | 0.8257 | 0.5392 | 1.264 | 0.9945 | 0.6621 | 1.494 | 0.8 |
| Biliary Obstruction | 0.8553 | 0.5400 | 1.355 | 0.8716 | 0.5541 | 1.371 | 0.8296 | 0.5256 | 1.309 | 0.9 |
| Ductopenia ≥30% | 1.299 | 0.7896 | 2.137 | 1.299 | 0.7896 | 2.137 | 1.299 | 0.7896 | 2.137 | 0.8 |
| Regenerative Hyperplasia | 0.9533 | 0.5406 | 1.681 | 0.9508 | 0.5459 | 1.656 | 0.6043 | 0.3222 | 1.133 | 0.4 |
| Chronic Rejection | 1.1477 | 0.6258 | 2.105 | .8969 | 0.4784 | 1.682 | 1.0444 | 0.5691 | 1.916 | 0.9 |
| De Novo Auto Immune Disease | 1.019 | 0.4237 | 2.449 | 1.425 | 0.6400 | 3.173 | 1.266 | 0.5543 | 2.892 | 0.8 |

| | | | | | | | | | | |
|---|---|---|---|---|---|---|---|---|---|---|
| Veno-Occlusive Disease | 1.435 | 0.5461 | 3.769 | 1.776 | 0.7087 | 4.453 | 1.463 | 0.5670 | 3.773 | 0.7 |
| Cirrhosis | 0.5764 | 0.1930 | 1.721 | 1.1200 | 0.4636 | 2.706 | 1.2493 | 0.5160 | 3.025 | 0.5 |
| Ductopenia ≥50% | 1.2082 | 0.4058 | 3.598 | 0.6309 | c0.1779 | 2.237 | 1.6069 | 0.5830 | 4.429 | 0.4 |
